# Substance use and associated factors among Adolescents’ during the Covid-19 pandemic in Eastern Ethiopia: A cross-sectional study

**DOI:** 10.1101/2022.03.29.22273151

**Authors:** Ebisa Zerihun, Kenasa Tesema

**Affiliations:** Department of Nursing, College of Health Science, Oda Bultum University, Chiro, Ethiopia; Department of Public Health, College of Medicine and Health Science, Haramaya University

**Keywords:** Adolescent, Covid-19 pandemic, Ethiopia, Substance use, West Hararge zone

## Abstract

**Introduction:** The effects of the COVID-19 pandemic have been dramatic and wide-reaching, affecting many more than those who become ill, including reports of increased substance use among adolescents may be due to various restrictions of social life that disrupted adolescents’ daily lives. However, up to now, no data is showing the extent of substance use among adolescents in the study area.

**Purpose:** This study aimed to assess the prevalence of substance use and associated factors during the Covid-19 pandemic among school adolescents of west hararghe, Eastern Ethiopia to guide possible intervention and public policy.

**Methods:** A School-based cross-sectional study design was conducted from 10 to 30, October 2021. A multi-stage sampling technique was used to select 788 students from ten public secondary schools in the West Hararge zone, Eastern Ethiopia. The data were collected using a self-administered pre-tested semi-structured questionnaire. Data was entered into Epidata version and then exported to SPSS version 26 software for further analysis. Descriptive analysis was done. Multi variables binary logistic regression was done and a p-value less than 0.05 were used to declare statistical significance.

**Results:** In this study, the response rate of 98.46% were complete fill the questionnaires. More than 1 in 2 of the adolescent students were self-reported substances (alcohol, khat (Catha edulis) and/or cigarette) users (prevalence = 58.6%) during the Covid-19 pandemic. Specifically, chewing khat (Catha edulis) (57.87%), followed by alcohol user (21.73%), Cigarette smoking (14.85%) and, hashish consumption (5.54%). Age of students, family history of drinking alcohol, availability of substances were factors positively associated with substance use. On the other hand, family management is negatively associated with substance use.

**Conclusions:** The prevalence of substance users among adolescents was dramatically increased during the covid-19 pandemic in the west hararghe zone, Ethiopia. Hence, the authors hoped that these findings provide preliminary insights for refining mental health and addiction policies that are targeted at adolescent and their parents in these settings and guidance for further research.

## 1. Introduction

According to World Health Organization (WHO), substance use was defined as taking one or more of the commonly used substances such as alcohol, cigarette, khat (Catha edulis), illegal drugs, prescription drugs and others in an individual’s lifetime to alter mood or behaviours(1). The most commonly used substances among adolescents vary depending on geographic locations worldwide. In Ethiopia, the most used substances among students include Khat (Catha edulis), cigarette, Alcohol like locally produced substances (araki, tella, and tej, beer, draft, etc)(2). Tella is a popular Ethiopian traditional beverage made from diverse ingredients or substrates such as barley, wheat, maize, millet, and sorghum(3). Araki is a colourless, clear, distilled, traditional alcoholic beverage in which fermented products are prepared in almost the same way as tella except that the fermentation mass in this case is more concentrated. Tej is a home processed and commercially available honey wine(4)

Substance use is a problem of public health importance across the world particularly among adolescents. It is a social problem that has spread and increased rapidly in educational institutions, especially among secondary school adolescents. This social problem is considered an issue of serious concern as it adversely affects the lives and performance of adolescents involved as well as the harmonious functioning of the entire structure of society(5). In today’s society, the use of substances is increasing in the adolescent population. This is due to several contributing factors, including peer influence, substance use within the family, media portrayals of substance abuse, and the use of negative coping mechanisms(6).

Before the COVID-19 pandemic in 2019, among North wollo, Ethiopian adolescents, the prevalence of substance use was 34.6%. Specifically, Khat (Catha edulis) chewer was 23.5%, Alcohol consumers were 23.5%, shisha smokers was 5.2%, and cigarette smokers were 3.3%.(7). Similarly, in 2019 among adolescents of Ambo town, central Ethiopia, the prevalence of substance users was 16% and about 10.5 % were Khat (Catha edulis) abusers, 2.6% were alcohol abusers, 3.8% were cigarette abusers and 1.01% were abused shisha(8). Likewise, in 2020 prevalence of substance use among adolescent students in the Kolfe-Keraniyo sub-city, Addis Ababa, Ethiopia, was 26.5%, while 16% drank alcohol, 9.6% smoked cigarettes, and 9.4% chewed khat (Catha edulis)(2).

Certainly, adolescence is a transitional phase of autonomy confirmation, peer relevance, and experimentation on life choices, which, combined with the financial and social perturbations during the pandemic, might predispose them to greater risks of substance use (9, 10). The disruption in daily routines for adolescents (through cancelled in-person classes, athletics, extracurricular activities and restrictions on public gatherings) during the Covid-19 pandemic might make them particularly vulnerable to substance use(11, 12). But less clear about the impact on substance use and possible factors for these effects.

The recent study explored the impact of physical distancing toward psychoactive substance usage, including their related factors among adolescents indicating changes in substance use behaviours during the pandemic could be unpredictable, as emotional distress, isolation, and unemployment drove the demand for substance use as a coping mechanism(13). Available research suggests the Covid-19 pandemic might increase depression among adolescents, leading to increased substance use, constituting an “epidemic hidden in a pandemic”(14). Similarly, the research found job loss during a population crisis was associated with increased psychological distress and substance use, even among persons still employed(15). In another way, reduced availability of substances, escalating prices, and financial limitations decreased substance usage(13).

Moreover, a correlational study suggested that the impact of the COVID-19 pandemic might differ across age and demographic groups, with adolescents being at particularly high risk for substance misuse. Due to several contributing factors include diminished peer interaction, greater proximity to parents, less participation in school-supported extracurricular activities, decreased availability of gathering spaces, less access to substance retailers, reduced exposure to substance use prevention programs, and heightened stress, anxiety, and boredom(16, 17).

Regarding the change of substance use in the general population including adolescents during the lockdown, two published studies to date found that alcohol and tobacco use increased in the general population but not use of other substances(18, 19). One study stated that individuals consumed slightly more alcohol and also smoked marginally more cigarettes compared to the period before the lockdown(18). Another study, that analyzed changes in alcohol drinking behaviour and tobacco smoking in the general population, found that 35.5% of the study population reported an increase in drinking during the lockdown(19).

Furthermore, at the beginning of the COVID-19 pandemic, the WHO warned that increased substance consumption during the lockdown can increase the prevalence of substance use disorders in the future(20). However, up to now no data shows the extent of substance use in the general population particularly among Ethiopian adolescents during the lockdown with regards despite the problem being omnipresent.

Therefore, this study was aimed to assess the extent of substance use and associated factors during the Covid-19 pandemic among secondary school adolescents of west Hararghe zone, Eastern Ethiopia. Because adolescent experiences different stressors during the pandemic that might mediate substance use in different ways, separate estimates are necessary to guide possible intervention and public policy. This finding will be provided baseline information for further researchers and health policymakers.

## 2. Methods

### Study Area and period

This study will be conducted in the Eastern part of Ethiopia countries, West hararghe zone administration of public secondary schools (from Grade 9^th^ to 12^th^). It is located 326 km (199 miles) East of Addis Ababa capital city of Ethiopia. There are about 50 public secondary schools in the West hararghe zone and there are about 54,260 students were enrolled in schools while as female 19737 (36.37%) and male 34523 (63.63%) students in 2021/2022 (West Hararghe Education bureau office September 02, 2021). The COVID-19 pandemic is considered to have begun after public schools across the nation closed and governors issued lockdown orders, starting from March 13, 2020, depending on respondents’ state of residence. Interviews began at local pandemic onset ended and school was reopened after lockdown from 10-30 /October 2021.

### Study design, and population

A school-Based Cross-Sectional Study design was employed. All randomly Selected Secondary school students who were enrolled in the 2021 /2022 academic year found west Hararghe zone during the study period were the study population. Students who have physical impairments (hearing, vision impairment) and are severely ill during the study period were excluded from this study.

### Sample size determination and sampling procedures

The sample size (n) required for this study was determined based on single population proportion formula with the assumption of 95 % C.I, 4 % margin of error and the overall prevalence of substance abuse of 26.5% from the previous study conducted among Secondary school students in Kolfe-Keraniyo sub-city, Addis Ababa, 2021(2). Finally, Considering, the 15% non-response rate and design effect of 1.5. The final sample size was **782**.

A multistage sampling technique was used to select the study participants. First, schools were divided into urban and rural, because the setting of the population is different. In the second stage, 10 schools (8 from rural and 2 from Urban) were selected randomly from 50 secondary schools (45 rural and 5 urban secondary schools) found in the zones. Then, further stratification was done based on the levels of study (grade 9^th^ to 12^th^). Finally, a simple random sampling technique was applied to select students in each grade of study from the list of students’ names in their respective grades. Students from each grade of the study were allocated proportionally to their class size. Eligible students were directly approached by the data collectors while they were at their respective classes in the morning time immediately after class attendance.

### Variables of the Study

The primary outcome variable of this study was substance use. Independent variables were

- Socio-demographic factors: Age, Sex, Ethnicity, Religion, Family drug use, Having pocket money, Family marital status
- Family-related Variables; Family substance use history, Relatives (same home) substance use history, Family Marital status
- Peer pressure factors; Drinking to satisfy friends, Not be seen as different from one’s group, Living with peers/friends currently substance use
- School-related factors (such as academic performance, school substance use regulation, and school substance controlling system)
- Environmental factors (such as availability of substances in residential areas, community norms on substance use, and availability of substance selling shops).

### Operational definition

Adolescence: The period from age 10 – 19 years

In this study, substance use was defined as taking one or more of the commonly used substances in Ethiopia, such as alcohol (including local liquors like tej, tella, areki), cigarette, khat (Catha edulis), shisha, cannabis, and others in an individual’s lifetime during the Covid-19 pandemic to alter mood or behaviours. Students who took one or more of the aforementioned commonly used substances within 30 days before the data collection period was taken as current substance users.

### Data collection tool and procedures

The questionnaire was developed after an extensive review of the literature and similar study tools used previously(7, 8, 16) by adapting to the purpose of the study. The questionnaire specifically constituted socio-demographic variables, substance use, and associated factors such as family, community, school, and environmental factors that influence the substance use of students. The data was collected using a pretested self-administered semi-structured questionnaire which was prepared in English and then translated to the local language (Afaan Oromo) which most students could understand from 10-30 /October 2021. The data collectors were Bachelor of Science degree (BSc) holders in Nursing who have guided the students to complete the questionnaire. The data collectors explain each question to the students to help them understand the questions well and fill their responses on the questionnaire. Facilitators were academic staff who were familiar with the specific school and facilitated the smooth running of the data collection process before and during the data collection period. The principal investigators have followed and controlled the overall data collection process, trained data collectors and facilitators. The Covid-19 preventive measures (wearing mask, physical distance, using alcohol, etc.) were maintained overall during data collection.

### Data quality management and analysis

A two days training were given for data collectors and facilitators. A pre-test was done on 5% of the total sample size outside of the study area. Data was entered into Epi-data version 3.1 and then exported to SPSS version 26 software for analysis. Descriptive statistics were computed to describe the characteristics of the participants. Binary variable logistic regression analysis was employed and variables at a p-value of less than 0.2 were candidates for multivariate analysis. Multivariate logistic regression analysis was carried out to control confounding variables and the association between independent and substance use was measured using AOR with 95% CI and P-value less than 0.05 was considered statistically significant. The Goodness of fit of the final logistic model will be tested. Finally, the result of this study was presented in graph, table and text according to the type of data.

### Ethical considerations

Ethical clearance was obtained from the ethical review committee of Oda Bultum University and an official letter was submitted to the West Hararghe educational office. A permission letter was also obtained from the West Hararghe education office and directed to each sampled school principal. Permission to conduct the research among students in the school was obtained from the school principal. After the purpose and the importance of the study were clearly explained, verbal informed consent was obtained from each participant and the information collected from each respondent was kept with complete confidentiality.

## 3. Results

### 3.1. Socio-demographic Characteristics of the students

From the total sample of 782 participants, students fill the questionnaire with completeness giving a response rate of 98.46%. Almost half of the participants 414 (53.77%) were male and 456 (59.22%) was Muslims. The mean age of students was 17.01 (SD ± 1.98years). The majority (83.24%) of surveyed students were from rural areas. Three hundred thirty-nine (43.79%) of student families attended secondary education while major sources of their family income were dependent on the farmer 211 (27.26%) and 558 (72.56%) of parents were providing pocket money for their child (see Table 1)

**Table 1:**
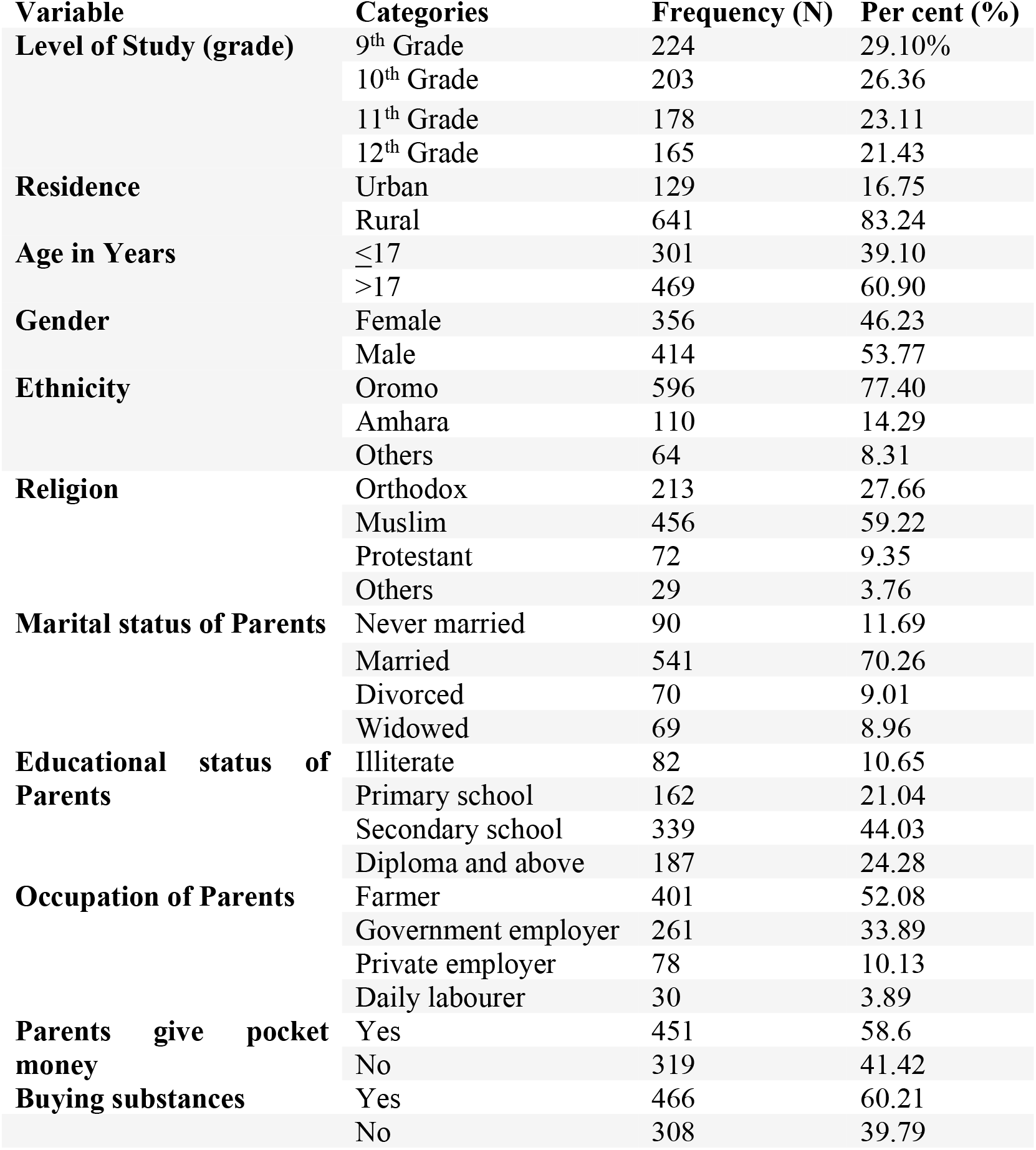
Socio-demographic characteristics of west hararghe zone secondary school students October 2021 (n=770)

### 3.2. Family-Related Variables

The overall score for family members using substances was 633 (82.96%). More than 459 (59.3%) of students reported that their parents have a history of drinking alcohol, while about 643 (83%) & 541(70.25%) families were chewed khat (Catha edulis) and smokers respectively. About 637 (82.3%) of their families have a history of behavioural problems. The majority of 610 (79.22%) students were reported that there has family management (follows up on the time coming home their child) (see Figure 1).

**Figure 1.**
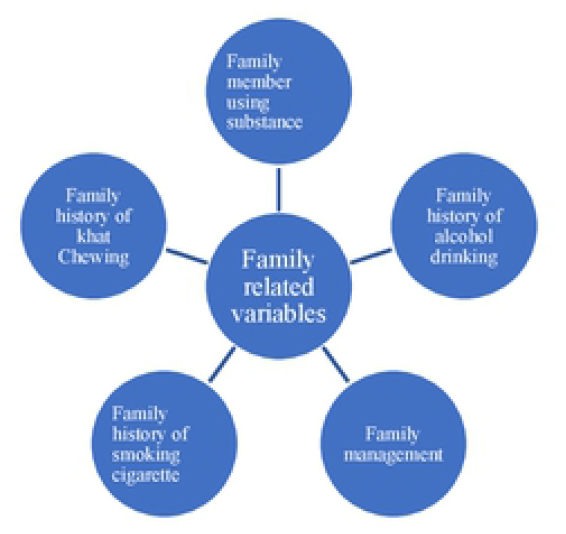
Family-related Variables among school adolescents during covid-19 in west hararghe, October 2021 (n=770)

### 3.3. Substances use of the students

More than 1 in 2 of the students were self-reported that they ever at least used substances (alcohol, khat (Catha edulis) and/or cigarette, others) during the Covid-19 pandemic. The over prevalence current substance use was 58.6% (95%CI = 55.09%, 62.10%) during Covid-19 pandemic. Regarding substance use, the respondents reported that they currently using the substance, of these 261(57.87%) chew khat (Catha edulis), 98 (21.73%) drink alcohol, 67 (14.85%) smoked cigarettes, and 25 (5.54%) took hashish (see Figure 2).

**Figure 2:**
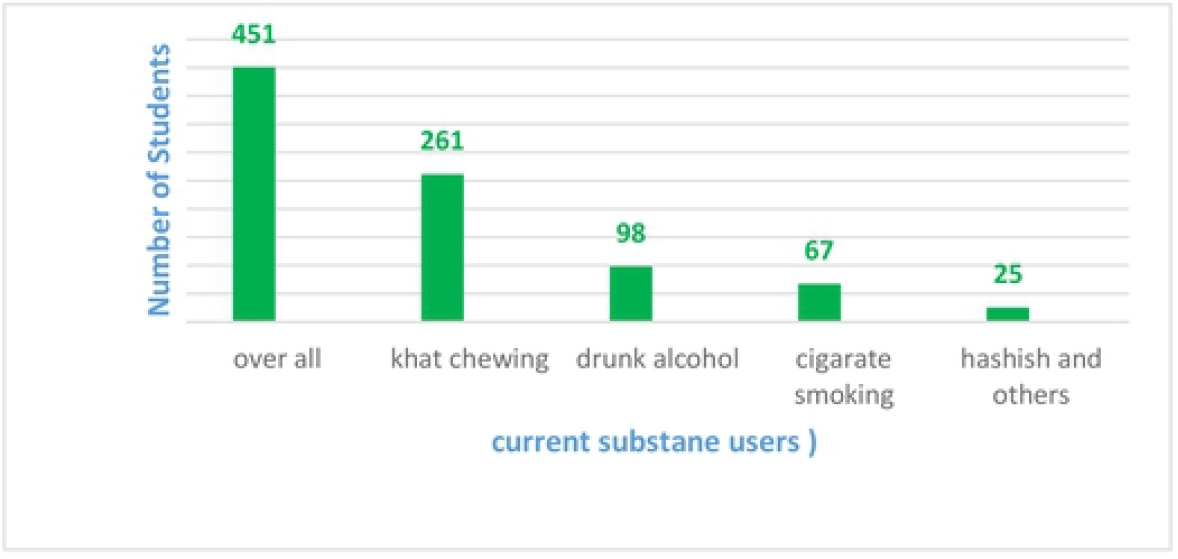
Current Substances Users among Secondary School Students in West Hararghe during Covid-19 October 2021 (n=770)

#### 3.3.1. Reasons for Substance Use

Different reasons were mentioned by students for the use of substances to relieve the stress of the covid 19 pandemic, to improves academic performance while schools re-opened, easily availability of substance retailing shops around school and in their residential areas and peer pressure reasons (friends’ delinquent behaviour, having friend use of substances, peer rewards, friends use of substances), etc. were reported the reasons facilitates the risk of using substance abuses.

#### 3.3.2. Protective factors from substance use

Study participants were asked about protecting reasons from substance use. Accordingly, 309 (40.13%) of respondents reported that their religion made them not use a substance. Similarly, 298 (38.7%) of respondents were found not using substances due to family control. Seventy-two (9.35%) of the respondents also mentioned other factors (like, because they did not want and believe substance use is harmful) which influenced them from using substances. The majority 543 (68.9%) of students agreed that adults in your neighbourhood think it is wrong for kids their age to use any substance and reported that their community has a disfavoring stand toward substance use. Five hundred seventy-four (74.16%) of the participants reported that their school has rules to control substance use, and mentioned reasons substance use controlling system in their school was ineffective.

### 3.4. Factors Associated with substances use

In the current study, Certain variables such as Age of students, Family history of drinking, & availability of substances in a residential area that act as a facilitator and having family management which acts as a barrier of substance use were associated with substance use among adolescents during the COVID-19 pandemic

The current study indicated that current substance use was statistically associated with the age of students. The odds of current substance use were about 2 times more likely to be higher among age greater than or equal to 17 years students compared with age less than 17 years of age (AOR = 1.85, 95%CI: 1.03, 3.34). Substance use among students was also significantly associated with the history of alcohol use in their family members. Students whose families have a history of drinking alcohol are about 3 times more likely exposed to substance use than those whose families do not drink (AOR = 2.62, 95% CI: 1.76, 3.88). Similarly, currently, the availability of substances in schools around and /or in the residential area increased the likelihood of substance use (AOR = 1.79, 95%CI: 1.22, 2.63).

Good family management was among the factors significantly associated with the current substance use of the students. Good family management is one of the protective factors identified. Students whose family follows up on the time coming home of their child reported about 60% lesser odds of current substance use as compared to their counterparts (AOR = 0.59, 95% CI: 0.36, 0.99) (see **Table 2**.)

**Table 2.**
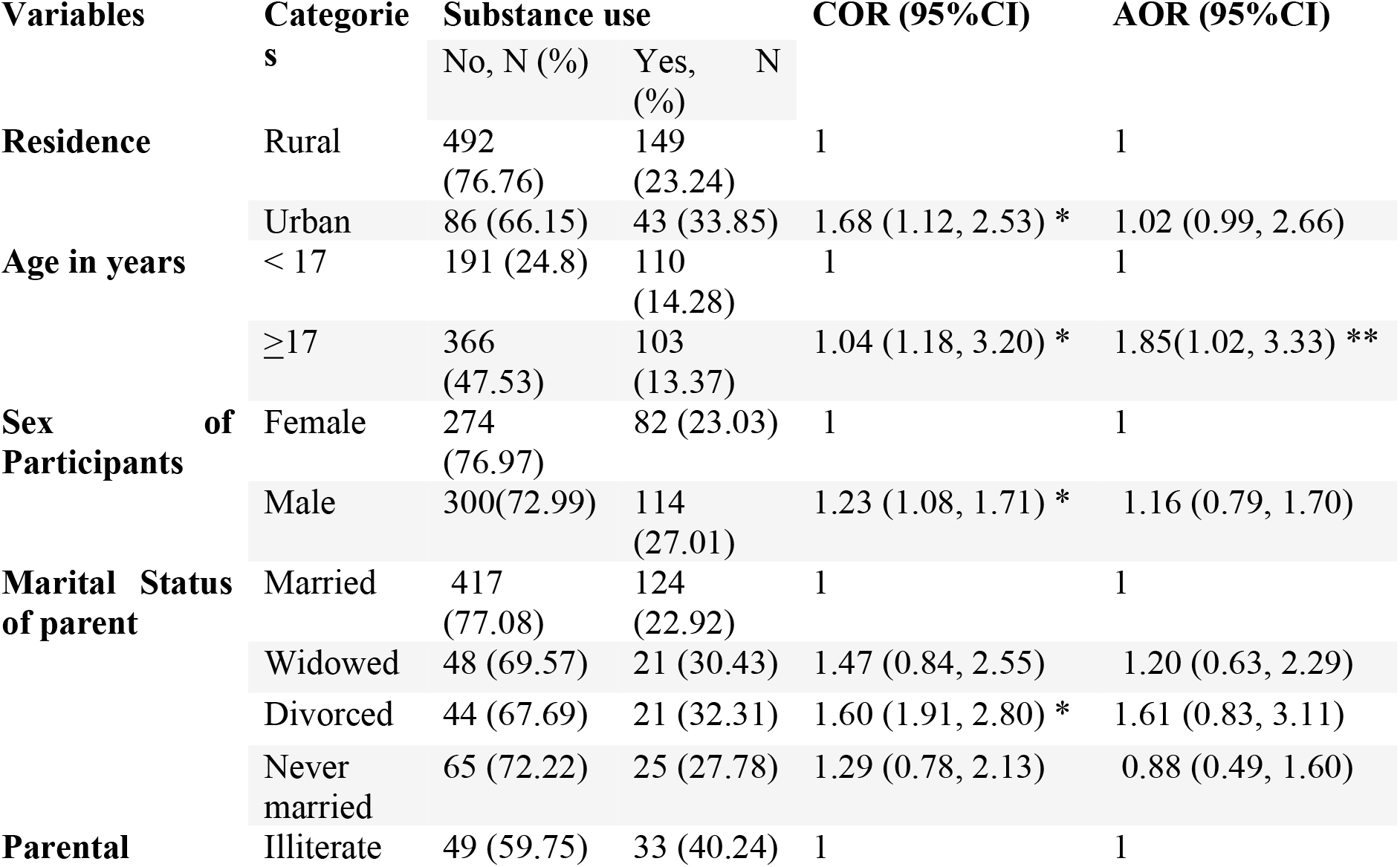
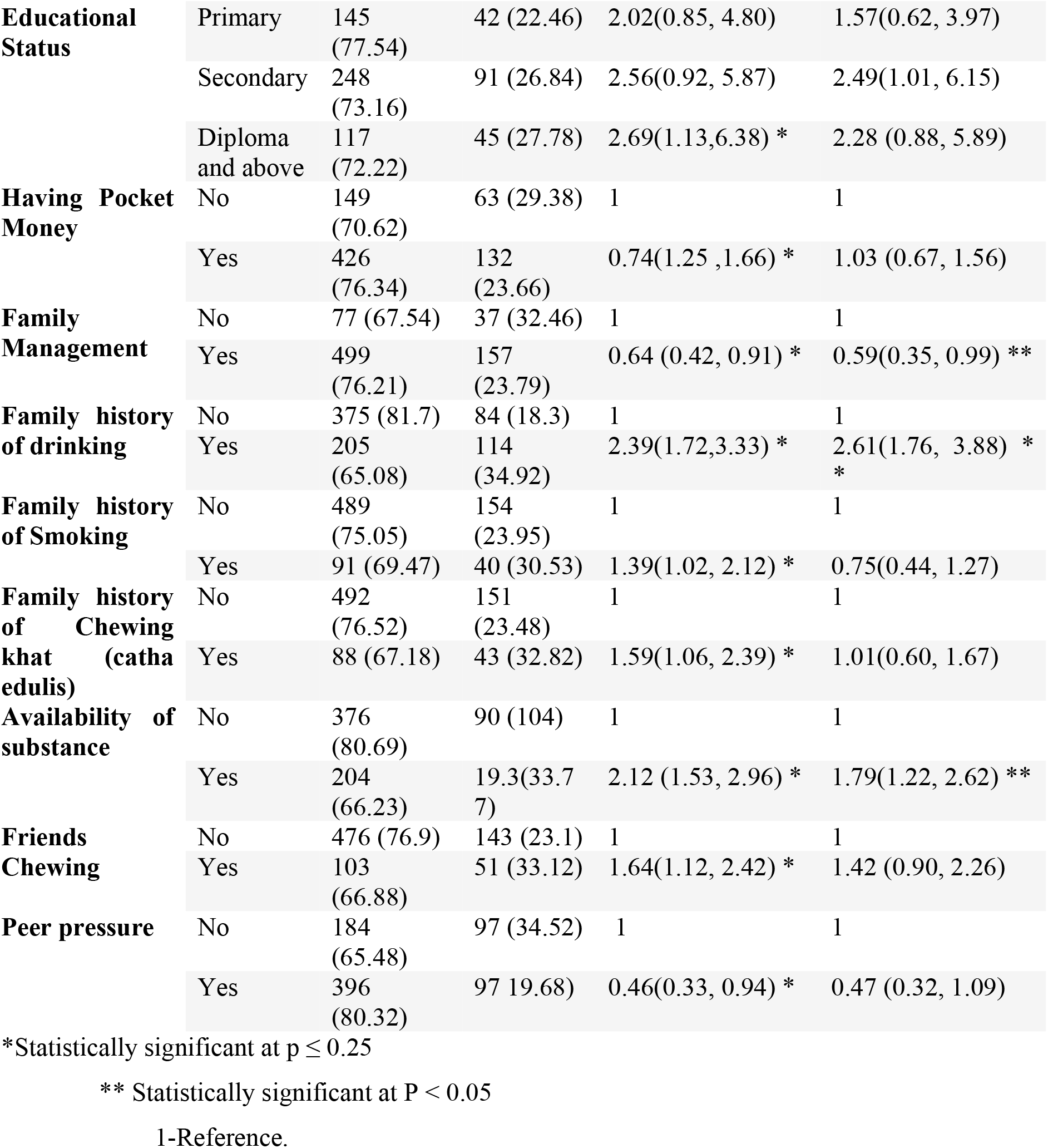
Factors Associated with Substances use among secondary School Students in west hararghe zone, October/2021 (n= 770)

## 4. Discussion

This study identified the overall prevalence of substance uses among secondary school adolescents of west Hararghe Eastern Eastern Ethiopia during the Cocid-19 pandemic was 58.6%. The rate of substance use differed for each type of substance, with the highest figure being chewing khat (Catha edulis) (57.87%), followed by alcohol user (21.73%), Cigarette smoking (14.85%) and, hashish consumption (5.54%). Overall, the difference might be chewing khat (Catha edulis) is the highest figure the fact that chewing khat (Catha edulis) in this study was conducted has been embedded in the social and cultural acceptability and also accessibility of khat (Catha edulis) in homestead lands also aggravate khat (Catha edulis) chewing among students(21).

This finding is higher than before the covid-19 pandemic conducted in a past different setting of the Ethiopian adolescent survey (2, 7, 8). Naturally, adolescence is a transitional phase of autonomy confirmation, peer relevance, and experimentation on life choices, which, combined with the financial and social perturbations during the pandemic, might predispose them to greater risks(9, 10). Additionally, brain development continues during the adolescence period; thus, teenagers tend to act impulsively without reflective thinking and are more vulnerable to addictive behaviours. The adolescent brain is also sensitive to the effect of psychoactive substances; therefore, it may damage the nervous system and affect brain functioning(22). These composite heightened vulnerabilities were reflected as higher substance use than in a past different setting of Ethiopian adolescent survey(2, 7, 8) and resonated in this study

This study identified that substance use is associated with the age of students. Students whose ages were greater than 17 years had more likelihood of substance use than less than 17 years. (AOR = 1.85, 95% CI 1.03, 3.34). This result is in agreement with the previous study conducted among Secondary School Adolescents in Nigeria Kagoro (23). Late adolescent age is the critical period in human life that need close supervision as students in this age category are eager to practice what they encounter.

In this study, having a family member history of drinking alcohol was associated with substance use among students. Students whose families drink alcohol have about 3 times more likely to use substances compared to those whose families do not drink (AOR = 2.62, 95% CI 1.76, 3.88). This finding was in line with a previous study conducted in woreta town, Bale zone, kolfe keranios Addis Ababa(24, 25). Parents have a huge role to play in terms of disciplining and leading by example since children learn by modelling and at such imitate their parents’ behaviour. Families who use a substance in front of their children became role models to their children and made children believe that substance use by adults is acceptable behaviour. Since there is a strong link between family and children, there might be adverse family-student experiences among Children and their family conduct. Therefore, the family should take care of their daily activities, so that their children will be protected against unhealthy lifestyles.

This study reported that the availability of substances in residential areas was significantly associated with increased substance use during the covid 19 pandemic (AOR = 1.79, 95%CI 1.22, 2.63). This finding was similar to previous studies conducted before the covid 19 pandemic (8, 26). This indicates the easy availability of substances to students might increase the risks of their consumption, thus, measures have to be taken to ensure that substance availability and accessibility to secondary school students is almost impossible

In this study, good family management is one of the protective factors identified. Students with have good family management (following up on the time coming home from their child) were less likely to use the substance compared to counterparts. This finding was similar to the study conducted in Nigeria(27). Families who were spent more time with their children and support increased the openness of children to share everything with their families and families therefore able to monitor and control unwanted behaviours of their children including substance use, thus parental supervision is necessary.

### Strengthen and limitations

The main strength of this study was being that the first study identified the extent of adolescent substance use during the COVID-19 pandemic in Ethiopia. However, there were some limitations in this study. First, some specifics of substance use attributes (e.g., history of use disorder, detailed categories, procurement sources, and adverse health effects) were not captured, thus requiring further research. Secondly, the study could not employ qualitative data, so it might not address more associated factors. Despite limitations, this study sheds important light on substance use trends and correlates among adolescents during the COVID-19 pandemic

## 5. Conclusion and recommendations

Most health policy attention has been placed on controlling the spread of the COVID-19 virus, but the current findings indicate a strong need also to address the substance use dramatically increased among all adolescents, more than half of the study participants were substance users. Khat (Catha edulis), Alcohol, and cigarette were widely used substances. Age of students, family history of drinking alcohol, availability of substance in a residential area, and having family management were identified factors significantly associated with substance use among secondary school students during the covid 19 pandemic.

Based on the findings of this study, the researcher made the following recommendations;

- Parents should always effort to monitor and keep a close check on their children, so they do not engage in substance abuse
- Parents should endeavour to show a good example to their children by not using the substance of abuse in their presence which could encourage them to start using the substance of abuse
- School-based program like substance-free clubs should be established in all secondary schools where drug/substance-related topics will be discussed which will enlighten and discourage students from substance abuse.
- Government should put restrictions on the category of people substances (alcohol, Khat (Catha edulis) and cigarette) can be traded based on age to reduce the accessibility of children to them and strengthen the school rules and policy on substance use
- Teachers should familiarize themselves with their students, so they can discover any anti-social behaviour among the students and provide immediate solutions to it
- Furthermore, the authors trusted that these findings provide preliminary insights for refining mental health and substance dependence policies and guidance for further research.
- Future research should examine adolescent substance use trends across time as the pandemic continues to affect daily life

## Data Availability

All relevant data are within the manuscript and corresponding authors

## Abbreviations

AOR: Adjusted Odds Ratio
CI: Confidence Interval
COVID-19: Corona Virus Diseases-19
SPSS: Statistical Package for the Social Sciences
SSA: Sub-Sahara African
WHO: World Health Organization

## Acknowledgement

Above all, we would like to heartfelt thanks to Oda Bultum University for giving us the opportunity of this work and our special thanks go to the West Hararghe Education Office for their genuine cooperation in providing us with updated students statistics. Last but, not least we would like to extend our gratitude to data collectors, facilitators and study participants whose efforts were crucial.

